# Cognitive reserve in a sample of Cuban older adults with Mild Cognitive Impairment

**DOI:** 10.1101/2021.07.31.21261435

**Authors:** Elizabeth Jiménez-Puig, Ana Cristina Baute-Abreu, Daniela Liz Valdés-Pérez, Yeisel Santana-Domínguez, Marina Jesús Añón-Villegas, Brayan Deivi Pérez-Leiva

## Abstract

**Introduction:** Aging is a natural process that occurs in all individuals. Due to the increase in aging figures worldwide, the diagnosis of mild cognitive impairment and other dementia syndromes has increased. An element of essential relevance in protecting against the risk of suffering from dementia is the cognitive reserve.

**Objective:** To indicate the levels of cognitive reserve in a sample of Cuban older adults with Mild Cognitive Impairment.

**Methods:** An exploratory study was carried out using questionnaires, with a cross-sectional, quantitative design. The sampling was non-probabilistic and intentional, obtaining a sample of 51 older adults with mild cognitive impairment. The information collection was carried out from the Cognitive Reserve Scale (CRS). Quantitative data was analyzed through frequency analysis.

**Results:** Most of the participants showed higher levels of cognitive reserve for carrying out activities of daily life and for relating to other people. The dimensions most affected were Formation / Information and Hobbies.

**Conclusions:** The research allowed a preliminary approach to the cognitive reserve of older adults. It would be interesting to conduct investigations of this nature in larger samples, including groups of healthy controls, which would allow comparison of results. In addition, it would be relevant to investigate the impact of institutionalization.

## Introduction

The increase in life expectancy has shown that the generational group of the elderly is expanding more and more.^(1)^ It is expected that by 2050 Cuba will be one of nations with the highest rate of population aging in the world.^(2)^

Mild Cognitive Impairment (MCI) has been conceptualized as a border line between normal aging and dementia. The cognitive decline that presents in MCI is not a part of normative aging, although it is not severe enough to diagnose dementia.^(3)^ Prevalence of MCI in population from 75 years of age is 3 to 20%, which represents an average of 8 to 60 people per 1000 in the year.^(4)^ In Cuba, prevalence have been found between 7.76 and 16.5% in people over 65 years of age and 40% in people aged 85 and over.^(5)^

However, there are some factors that slow down or protect the brain against deterioration. Cognitive reserve (CR) is known as the ability of our body to resist brain deterioration without the presence of symptoms. The CR theory states that people with a higher CR have a lower risk of suffering from a degenerative pathological process in cognitive impairment during old age, although this normal impairment also decreases.^(6)^ The cognitive reserve not only allows an improvement in mental health, but also in physical. There are some activities that increases and maintain CR, such as education; leisure activities; cultural, social and cognitive activities. All of them requires the implementation of cognitive processes and can contribute to the development of the cognitive reserve.^(7)^ CR is accumulated through a life spam, therefore, it is present in healthy people and in people with some levels of deterioration.^(6)^ That is why the present study aims to indicate the levels of CR in a sample of Cuban older adults with MCI.

## Methods

An exploratory study was carried out using questionnaires, with a cross-sectional, quantitative design. The population was institutionalized older adults, from Villa Clara and Cienfuegos provinces (Cuba). The study was carried out from November 2019 to March 2020. The sampling was non-probabilistic and intentional, through selection criteria.

### Inclusion criteria

- To sign up the Informed Consent.
- Older adults with an age equal to or greater than 55 years.
- To obtain less than 90 points on the Addenbrooke’s Cognitive Examination III (ACE-III).

### Exclusion criteria

- Presence of clinical signs of anxiety (score greater than 40 points on the Zung Anxiety Test).
- Presence of clinical signs of depression (score greater than 4 points on the Geriatric Depression Scale).
- Illiteracy.
- Presence of visual, auditory or motor perceptual disorders.
- The use of psychoactive drugs that affect the level of wakefulness.

### Exit criteria

- Explicit desire to abandon the research.
- Incomplete sessions.
- Appearance of physical or mental at the moment of investigation.

The sample was made up of 51 Cuban older adults with MCI (table 1).

**Table 1.**
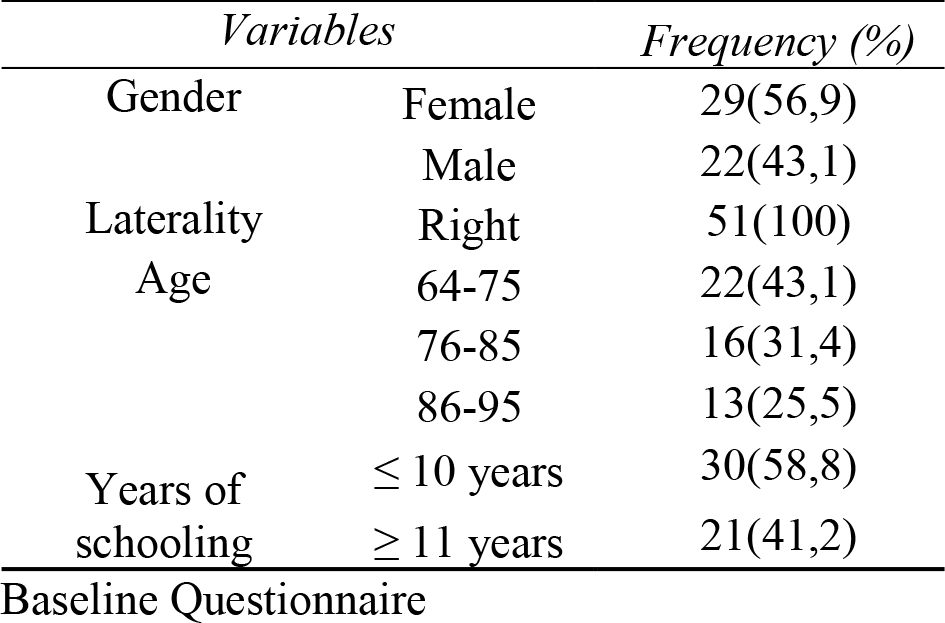
Sociodemographic variables

| *Variables* |  | *Frequency (%)* |
| --- | --- | --- |
| Gender | Female | 29(56,9) |
|  | Male | 22(43,1) |
| Laterality | Right | 51(100) |
| Age | 64-75 | 22(43,1) |
|  | 76-85 | 16(31,4) |
|  | 86-95 | 13(25,5) |
| Years of schooling | ≤ 10 years | 30(58,8) |
|  | ≥ 11 years | 21(41,2) |

Baseline Questionnaire

Cognitive Reserve Scale (CRS): The objective it to estimate the cognitive reserve of an individual through the collection of information about their entire adult life.^(8)^ It is divided into four facets: activities of daily living, formation-information, hobbies, and social life. The response options present in a Likert scale. It is estimated that the higher the scores, the higher the cognitive reserve index. In this study, the total scores were divided taking as a reference the mean of total scores obtained. The reliability index of the instrument was calculated through Cronbach’s Alpha for its use in the present investigation. It gave a value of α = .92.

### Procedures

Prior to the beginning of the evaluation sessions, the corresponding consents were requested from the directors of each of the institutions in which the research was carried out. The research was approved by the Senior Citizen Care Program and by the Ethics Committee of Central University Marta Abreu of Las Villas.

The objectives of the research were explained to the subjects and their consent was obtained to participate. The ACE III, the Geriatric Depression Scale and the Zung Anxiety Test were applied. Subsequently, the Cognitive Reserve Scale was applied.

### Data analysis

The data were analyzed using the statistical processor SPSS (Statistical Package for the Social Sciences) in its version 25.0 for Windows. They were analyzed through frequency analyzes.

## Results

In the facet of Activities of Daily Living, no great difference was reflected in the levels of CR, so it can be affirmed that the percentage in this case was quite balanced. In the item related to Social Life, the results were similar. On the other hand, lower levels of CR were found in the Formation/Information facet (74.5 % of the participants). On the Hobbies facet, the entire sample showed lower levels of CR (table 2).

**Table 2.**
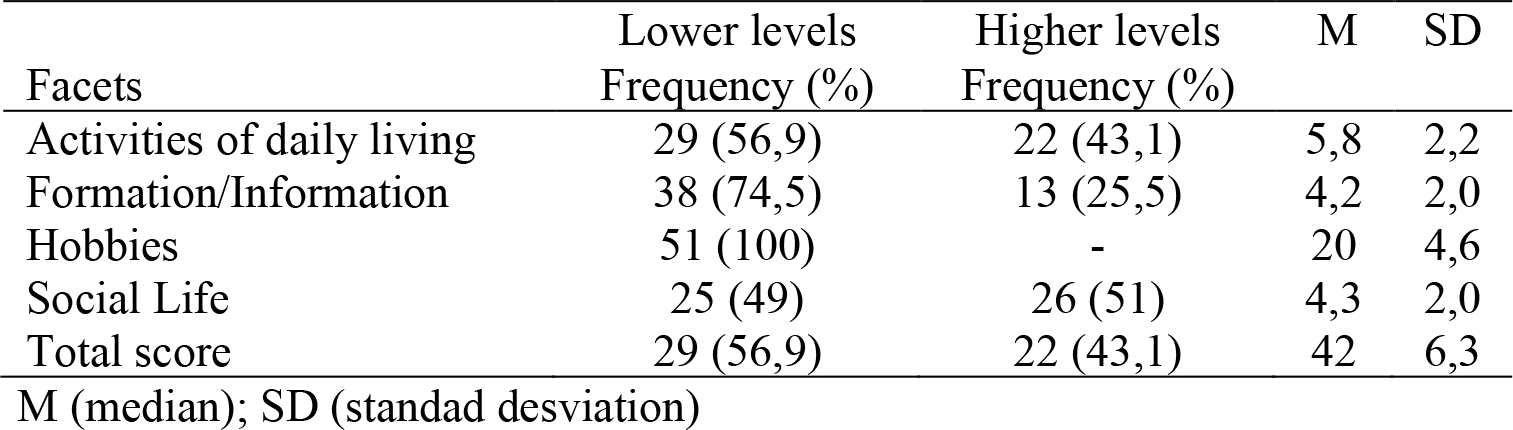
Cognitive Reserve Levels

## Discussion

The cognitive reserve accumulates throughout life and includes both innate and acquired elements, such as schooling, occupation, the performance of cognitively challenging activities and leisure in adulthood and older adulthood, the IQ, as well as measures of different cognitive functions. Among the elements acquired are the reading ability, hobbies, and physical activity.(9) Among the predictive factors of cognitive reserve, which are created throughout the life to cope with deterioration in old age, education and intellectual capacity, occupational complexity and leisure activities have been described, such as crafts, arts, music, physical activity, reading, intellectual games, among other hobbies.^(6)^

Over half of our sample can perform household chores and take care of their personal affairs. They can perform basic activities of daily living and they are functionally independendent. Almost half of the individuals evaluated did not have difficulty visiting their friends or relating to people from other generations; issue that can also be analyzed as a protective factor throughout life. In other words, the Social Life dimension scored high in almost all the life spheres to which the test refers. This result is relevant since the level of cognitive reserve can be greatly enhanced with the performance of activities of this nature. There is growing evidence that a lifestyle characterized by greater involvement in leisure activities of a social nature is associated with a slower cognitive decline in the elderly. It also reduces the risk of dementia because the creation of efficient cognitive networks provides a cognitive reserve that contributes to delaying the clinical manifestation. Developing a socially committed lifestyle favors the maintenance of verbal intelligence in elderly people.^(10)^

Nevertheless, in our sample, there were lower levels of CR in educational activities and hobbies. This first element could be related to the number of years of schooling reported by older adults. Education has been the factor most strongly indicated as a predictor of reserve cognitive capacity; intellectual capacity is also considered as a predictive factor as robust as education. However, education is presented as a promoter and maintenance marker of high intellectual performance, so that intellectual capacity does not seem to be able to explain by itself cognitive reserve.^(11)^

On the other hand, it has been studied the impact of institutionalization in the elderly. This could be an important factor that attempt against CR if it is not properly worked. The institutionalized older adult is characterized by the loss of the social and family affective bond, which is produced by the convergence of one or more factors such as, distance from personal connections, loss of the work role, emotional deprivation, few family and friends ties, widowhood, financial problems, feelings of worthlessness, and physical or mental health problems.^(12)^ Institutionalization can accelerate cognitive decline, increases dependency, feelings of loneliness and handicap.

The authors acknowledge the importance of the present study; however, some limitations are recognized. In essence, the exploratory scope of the investigation and the sample size.

It would be interesting to conduct investigations of this nature in larger samples, including groups of healthy controls, which would allow comparison of results. In addition, it would be relevant to investigate the impact of institutionalization.

## Data Availability

None

